# Seroprevalence of *Brucella* antibodies among vaccine manufacturing workers in contact with the *Brucella melitensis* Rev.1 vaccine strain

**DOI:** 10.1101/2021.12.02.21267183

**Authors:** Manuel Vives-Soto, Amparo Puerta-García, José-Luis Pereira, Esteban Rodríguez-Sánchez, Javier Solera

## Abstract

**Background:** Cattle vaccination remains an essential measure for the control of brucellosis. Strict preventive measures are applied to protect vaccine manufacturing workers (VMW) employed in processing these live attenuated vaccines. We analyzed the serological responses of VMW in contact with the *Brucella melitensis* Rev.1 strain.

**Methods:** We conducted an observational study of a cohort of VMW in a Spanish biopharmaceutical company, a leader in manufacturing veterinary products. The results of the *Brucella* serological tests carried out on these workers between 2012 and 2019 were reviewed, as well as demographic data, length of time in the company (seniority), and level of exposure. Multivariate analysis was performed with the logistic regression test.

**Results:** Of the 115 VMW studied, 47 (41%) showed positive Rose Bengal tests during company check-ups. Exposure levels were correlated with seropositivity, with an adjusted OR of 6.6 (95% CI: 2.1-20.3) for the high exposure and 2.0 (95% CI: 0.6-6.7) for the medium exposure groups. Sixteen (34%) seropositive VMW demonstrated an acute serologic pattern of IgG and IgM antibodies seropositivization, while 31 (66%) manifested a chronic serologic pattern of constant or intermittent positive IgG antibodies with persistently negative IgM antibodies. Seniority was inversely associated with the acute pattern: adjusted OR of 0.88 (95% CI: 0.79-0.97) for each year added. No seropositive VMW showed evidence of active brucellosis during follow up.

**Conclusion:** Despite strict safety measures, a percentage of VMW were exposed to the Rev.1 strain. Exposure levels were correlated with seropositivity. None of them developed symptomatic infection during follow-up. Two different serological patterns were observed: an acute IgM-positive pattern or a chronic IgM-negative pattern. Seniority was associated with the chronic pattern.

## INTRODUCTION

Human brucellosis is one of the most common zoonotic infections worldwide. Although the number of cases of brucellosis may be decreasing in the world, the continued presence of the disease in some endemic areas and the potential use of *Brucella* species as an agent of bioterrorism, make brucellosis a major public health hazard with important sanitary and economic repercussions (1). *Brucella* is a facultative intracellular microorganism capable of surviving and multiplying within the cells of the reticuloendothelial system, evading immune defense mechanisms and sometimes causing chronic or persistent infections. It enters the susceptible host via the oral, nasal, conjunctival, or genital routes, and triggers an innate and adaptive immune response. Although both antibody- and cell-mediated immune responses are diagnostically useful, antibodies are easier to measure quantitatively; therefore, serologic tests continue to be used to diagnose this infection, given the risk and difficulty involved in obtaining positive cultures (2).

Currently, cattle vaccination remains an essential measure for the control of this zoonosis, and only live attenuated vaccines (also called agglutinogen vaccines) have shown efficacy in preventing infection in cattle. The strains currently used in most countries for the control of bovine brucellosis are *Brucella abortus* S19 and *Brucella abortus* RB51, while the strain used for small ruminants is *Brucella melitensis* Rev.1 (3). These strains are capable of establishing limited infection in cattle, mimicking the natural infection process by wild strains, and thus conferring protection. However, these vaccines are not used in humans due to the high risk of developing acute brucellosis (4). Therefore, in order to protect vaccine manufacturing workers (VMW) from being exposed to these *Brucella* strains during the vaccine manufacturing process, strict safety measures in compliance with the Biosafety in Microbiological and Biomedical Laboratories (BMBL) standards are currently in place, involving: a) at least class II Biological Safety Cabinets (BSCs), b) proper personal protective equipment (PPE) and c) use of primary and secondary barriers (5). Notwithstanding, a certain likelihood of exposure persists, and companies keep an active watch on these VMW (6).

The goal of the present study was to determine the likelihood of seroconversion in VMW producing the *Brucella melitensis* Rev.1 vaccine strain, as well as observe the different patterns of serological response to the strain.

## MATERIAL AND METHODS

We conducted an observational study of a cohort of VMW at CZ Vaccines, a Spanish biopharmaceutical company with international projection, belonging to the Zendal group. Located in O Porriño (Pontevedra, Spain), it is a leader in the production and distribution of cattle vaccines, including the *Brucella melitensi*s Rev.1 strain. An annual serological check-up, using the Rose Bengal test, is systematically carried out on all workers in the company potentially exposed to this strain. If positive, IgG and IgM levels are determined by Enzyme Immunoassay (EIA). Using EIA, an index ≥1.1 is considered positive and an index ≤0.9, negative. To determine whether there is active infection, Polimerase Chain Reaction (PCR) is performed by rt-PCR, and *Brucella* spp. is sequenced by molecular hybridization.

The Rose Bengal (RB) test consists of slide agglutination with *Brucella* suspended in lactate buffer, colored with Rose Bengal. It is a very useful and rapid initial or screening test, positive in 99% of patients who suffer from brucellosis or who have had direct contact with the germ (6).

Enzyme immunoassay tests can detect specific immunoglobulins with sensitivities ranging from 93 to 97% and a specificity of 98%. A good correlation has been found between IgM levels and serum agglutination titers in tube or wellplate, as well as between IgGs and the Coombs and complement fixation tests (7). During the acute phase of brucellosis, there is a classic antibody response with increased IgM and IgG. Throughout the course of the disease, the IgM titer decreases, but can frecuently be detected up to 8-10 months following initial infection. Likewise, although IgG antibody titers usually decrease after onset of effective antibiotic treatment, significant titers may persist for several months or even years, despite cure (2). This fact makes it difficult to differentiate between active infection, history of brucellosis or a non-clinically relevant immune memory resulting from repeated exposure to the etiologic agent. Therefore, high antibody titers can lead to unnecessary long-term antibiotic therapy (8).

The Polymerase Chain Reaction (PCR) technique detects *Brucella* DNA with a very high sensitivity and specificity, allowing patients with suspected active brucellosis to be diagnosed with certainty (9).

### Design

VMW were categorized into three groups: high (bacteria culture management), medium (freeze-drying and packaging jobs) and low (labeling and other administrative tasks) levels of exposure. Workers who had rotated through various jobs were included in the group corresponding to the highest exposure. We analyzed the results of the Rose Bengal serological screening tests for *Brucella* carried out between 2012 and 2019 on VMW potentially exposed to the Rev.1 strain, as well as their demographic data and seniority in the company. All VMW with positive Rose Bengal tests were included in the EIA analysis of IgG and IgM serological response patterns and were followed for at least 2 years. All samples were analyzed in a central reference laboratory with commercial Kits: Biorad for the RB test, Vircell for IgG and IgM EIA, and an in-house method with TIB-Molbiol primers for the rt-PCR test.

### Statistical analysis

The statistical program SPSS Statistics v.21 was used. The qualitative variables were compared using the Chi-square test, with post-hoc analysis of the adjusted residuals when there were more than two categories. Quantitative variables were described as medians (interquartile range) and were compared using the Kruskal-Wallis test. For the multivariate analysis, logistic regression with the retrograde conditional method was used. A bilateral p ≤0.05 was considered significant.

## RESULTS

The study period was 8 years (2012 - 2019). 115 VMW were included: 47men (41%) and 68 women (59%), with a mean age of 42 years (range 23 - 66 years). The mean seniority in the company was 12 years (range 1 - 51 years). The level of exposure to the Rev.1 strain was high in 50 (43%), medium in 38 (33%) and low in 27 (24%) VMW.

To assess the likelihood of seroconversion in VMW in contact with the Rev. 1 strain, the Rose Bengal test was used as a reference: 47 became seropositive (41%) at some point during the study period and 68 remained seronegative (59%). **Table 1** summarizes the results of the bivariate analysis. Seropositivity was associated with both level of exposure and length of time in the company. However, in the multivariate analysis, only level of exposure was statistically significant, with adjusted odds ratios (OR) of 6.6 (95% CI: 2.1 – 20.3) for high exposure and 2.03 (95% CI: 0.6 – 6.7) for medium exposure when compared with the low exposure subgroup. No seropositive VMW developed symptoms of active brucellosis. *Brucella* spp. PCR was performed in whole blood on 6 VMW with seroconversion and positive IgM, with negative results in each case.

**Table-1:** Analysis of the risk factors for seroconversion of the 115 VMW.

|  | RB+ (n=47) | RB- (n=68) | P |
| --- | --- | --- | --- |
| Male sex | 19 (40%) | 28 (41%) | 0.963 |
| Age (years) | 43 (37-47) | 38 (34 - 44) | 0.066 |
| Seniority (years) | 13 (4 - 19) | 7 (2 - 17) | 0.041 |
| Level of exposure: |  |  | 0.001 |
| High | 30 (64%) | 20 (29%) |  |
| Medium | 12 (25%) | 26 (38%) |  |
| Low | 5(11%) | 22 (32%) |  |

VMW= vaccine manufacturing workers. RB+ = positive Rose Bengal test. RB- = negative Rose Bengal test. Quantitative variables: median (interquartile range). Qualitative variables: n (%).

In the 47 seropositive VMW, two different patterns of serological response were defined: an “acute” pattern of IgG and IgM seropositivization (pattern 1, n=16) and a “chronic” pattern of persistently negative IgM with positive and negative IgG determinations (pattern 2, n=31). Although most of the IgM positive VMW showed IgM negativization after 1 year (n=12), four VMW had persistent positive IgM for two or more years. **Table 2** summarizes the results of the bivariate analysis. Positive IgM patterns were associated with the youngest age group and inversely correlated with time spent in the company (“seniority”); however, in the multivariate analysis, only seniority was significantly linked to the pattern with an adjusted OR of 0.88 (95% CI: 0.79-0.97) for each extra year in the company.

**Table-2:** Factors associated with the serological patterns of the 47 seropositive VMW.

|  | IgM positive<br>(n=16) | IgM negative<br>(n=31) | P |
| --- | --- | --- | --- |
| Male sex | 8 (42%) | 9 (39%) | 0.739 |
| Age (years) | 36 (33-43) | 44 (41-50) | 0.002 |
| Seniority (years) | 6 (3-14) | 15 (9-21) | 0.006 |
| Level of exposure: |  |  | 0.738 |
| High | 9 (56%) | 21 (68%) |  |
| Medium | 5 (31%) | 7 (22%) |  |
| Low | 2 (13%) | 3 (10%) |  |

VMW= vaccine manufacturing workers. Quantitative variables: median (interquartile range). Qualitative variables: n (%).

## DISCUSSION

In this study, we analyzed the human serological response over time in a cohort of vaccine manufacturing workers (VMW) exposed to the attenuated *Brucella melitensis* Rev 1 vaccine strain. Despite their lesser virulence, these attenuated strains occasionally infect veterinarians due to accidental inoculation or splashes on mucous membranes (6,10) Additionally, brucellosis can be acquired by inhalation (11–14), and vaccine strains are also capable of airborne transmission (15). Therefore, there is a risk of becoming infected from occupational exposure to these strains. However, to the best of our knowledge, a systematic analysis of the serological evolution over time in a cohort of exposed VMW has not been conducted so far. Although none of the VMW in our study developed symptomatic brucellosis, seropositivity was observed in 41% of the individuals examined, indicating asymptomatic infection with the Rev.1 strain, despite strict safety measures. This seropositivity was significantly associated with a higher level of exposure to the Rev.1 strain. The fact that none of the VMW studied developed the disease could be explained by the lower virulence of the Rev.1 strain and / or by a smaller bacterial inoculum due to safety measures. Moreover, the seroconversion of these VMW may have actually protected them from contracting brucellosis, as has been previously described (14).

In 1987, Ollé-Goig et al. described an airborne-acquired outbreak of brucellosis in 28 workers who were cultivating the Rev.1 strain at a manufacturing plant for veterinary biologic products in Gerona (Spain) (15). The investigation included 172 workers, of which 28 were identified as having acute brucellosis, 20 had chronic brucellosis, 106 were infection-free, and 18 had no clear diagnosis. The cause of infection was a failure in the ventilation system, which resulted in an outbreak of brucellosis in nearby workers. Cases of accidental exposure to other vaccine strains have also been reported. Ashford et al. (2004) published findings from the Centers for Disease Control and Prevention (CDC) regarding a passive surveillance system for accidental inoculation with the RB51 vaccine in the United States (16). Reports were received from 26 individuals accidentally exposed during animal vaccination, but only one of them had a positive culture, which was taken from the cutaneous injection site. The authors concluded that it remains undetermined whether the RB51vaccine can cause systemic brucellosis in humans. However, Wallach et al. (2008) evaluated the pathological consequences of exposure to the vaccine strain *Brucella abortus* S19 in 30 employees from vaccine manufacturing plants, and active brucellosis was diagnosed in 21 of them (17). Only five of these VMW recalled an accidental exposure, indicating that employees from laboratories producing the S19 vaccine are at risk of exposure to *Brucella abortus* and may become infected by this strain. In 2018, Xie et al published a meta-analysis of 27 papers, reviewing the adverse effects associated with the three licensed brucellosis vaccines: S19, Rev1 and RB51 (18). The classification analysis showed that adverse effects in animals were found mainly in the immune and reproductive systems, while human adverse effects usually involved behavioral and neurological systems. Therefore, the risk of developing disease from the attenuated vaccine strains and the importance of taking extreme security measures cannot be minimized.

Regarding our study, two different patterns of serological response were identified in seropositive employees: an acute pattern of IgG and IgM seropositivization, and a chronic pattern of persistently negative IgM with positive and negative IgG determinations. These patterns were correlated with length of service: VMW who had been in the company for the shortest period were the most likely to be positive for IgM, and a small subgroup of them remained IgM positive over time (2 or more years), an exceptional fact in the literature. Although false positives for IgM against *Brucella* have been described in asymptomatic subjects (19), in all these cases the IgG was negative, unlike the subjects in our series. On the other hand, false-negative IgM results might be obtained due to an excess of IgG antibodies, and false-positive results due to the presence of rheumatoid factor (2). It is noteworthy that of the 4 subjects with persistently positive IgM, 3 belonged to the intermediate exposure group and 1 to the high risk group. In any case, it is clear that the presence of positive IgM does not indicate disease nor does it warrant antibiotic treatment.

Antibody titers usually decline after the initial infection, but significant titers may persist for several months or even years, despite negative blood cultures (2). This fact complicates the differentiation between active infection, history of brucellosis or a non-clinically relevant immune memory as a result of repeated exposure to the etiologic agent. Raised titers may consequently lead to unnecessary antibiotic therapy. Therefore, the sole detection of anti-*Brucella* antibodies does not provide evidence for the presence of the pathogen. High titers during follow-up are often related to high titers during the initial infection and are not always a sign of *Brucella* sickness, chronic disease or relapse. Particularly in endemic regions, a large proportion of the population may have persistent, specific antibodies due to continuous exposure to *Brucella*. Furthermore, relapse is characterized by a second peak of anti-*Brucella* IgG and IgA, but not by IgM immunoglobulins (8).

PCR has been successfully used to detect chronic brucellosis in patients with non-specific symptoms (9). In our study, the 6 patients who underwent PCR in blood had a negative result, despite presenting positive IgM seroconversion. This fact underscores the absence of active brucellosis. In a study carried out in our Hospital in 2010, *Brucella melitensis* DNA was analyzed in blood, serum and other biological samples in a cohort of 38 subjects with a previous history of brucellosis, diagnosed between 3 and 33 years earlier (20). The results emphasized the usefulness of PCR in identifying patients with brucellosis.

Our study suffers from some limitations. Firstly, follow-up times for the different VMW varied, as some left the company, while others were incorporated throughout the study. Secondly, exposure intensity for some VMW also varied over time. Additionally, the accuracy of the IgG and IgM serological patterns observed might have been affected by any false negatives results of the Rose Bengal screening test. Finally, the median length of service in the company was 12 years; therefore, we cannot know with certainty the long term health repercussions of seroconversion.

In conclusion, our study shows that, despite safety measures, a percentage of vaccine manufacturing workers develop immunity to the Rev.1 strain, a fact that is correlated with level of exposure. Secondly, it is noteworthy that none of them had evidence of active infection during follow-up. Finally, we identified different serological response patterns which were correlated with seniority in the company and, therefore, with an immune response to repeated exposures to the vaccine strain.

## Data Availability

All data produced in the present study are available upon reasonable request to the authors

## NOTES

### Author Declarations

We confirm that all relevant ethical guidelines were followed, and any necessary IRB and/or ethics committee approvals were obtained.

The details of the IRB/oversight body that provided approval or exemption for the research described are given below: Ethics Committee for Research with Medications of Galicia (CEImG), registration code 2021/422.

We followed all appropriate research reporting guidelines and uploaded the relevant EQUATOR Network research reporting checklists and other pertinent material as supplementary files, where applicable.

### Availability of all data

All data produced in the present study are available upon reasonable request to the authors.

### Author contributions

Conceptualization and coordination: ERS, JS; Data acquisition and anonymization: JLP; Data preparation and formal analysis: MVS, APG; Research: MVS, APG, JS; Writing – original draft preparation: MVS, APG, JS; Supervision: ERS, JS.

## Acknowledgments

The authors thank Alexandra Salewski Msc for expert English review of the manuscript, and Carlos de Cabo PhD for preparation of the edition and presentation of the work for its publication.

## BIBLIOGRAPHY

(1) Dean A, Crump L, Greter H, Schelling E, Zinsstag J. Global burden of human brucellosis: a systematic review of disease frequency. PloS NeglTrop Di 2012; 6 (10): e1865.

(2) Dahouk SA, Nöckler K. Implications of laboratory diagnosis on brucellosis therapy. Expert Rev Anti Infect Ther 2011; 9: 833–845.

(3) Carvalho TF, Haddad JP, Paixão TA, Santos RL. Meta-Analysis and advancement of Brucellosis vaccinology. Plos One 2016; 11: e0166582.

(4) Pink WW, Hall-III JW, Finstad J, Mallet E. Immunization with viable Brucella organisms: results of a safety test in humans.Bull World Health Organ 1962; 26: 409–419.

(5) Centers for Disease Control and Prevention. National Center for Emerging and Zoonotic Infectious Diseases. Brucellosis Reference Guide: Exposures, Testing, and Prevention. Updated February 2017. https://www.cdc.gov/brucellosis/pdf/brucellosi-reference-guide.pdf

(6) Berkelman RL. Human illness associated with use of veterinary vaccines. Clin Infect Dis 2003; 37: 407–414.

(7) Araj GF. Update on laboratory diagnosis of human brucellosis. Int J Antimicrob Agents 2010; 36 Suppl 1:S12–17.

(8) Solera J. Update on brucellosis: therapeutic challenges. Int J Antimicrob Agents. 2010; 36 Suppl 1:S18–20.

(9) Romero C, Gamazo C, Pardo M, López-Goñi I. Specific detection of Brucella DNA by PCR. J Clin Microbiol 1995; 33: 615–617.

(10) Blasco JM. Brucella melitensis Rev-1 vaccine as a cause of human brucellosis. Lancet 1993; 242: 805.

(11) Rodríguez ML, Pousa A, Pons C, Larrosa A, Sánchez LP, Martínez F. Brucellosis as an occupational disease: study of an airborne transmission outbreak in a slaughterhouse. Rev Esp Salud Pública 2001; 75:159–170.

(12) Buchanan TM, Faber LC, Feldman RA. Brucellosis in the United States, 1960-1972. An abattoir-associated disease. Part I. Clinical features and therapy. Medicine (Baltimore). 1974 Nov; 53(6): 403–13. doi:10.1097/00005792-197411000-00001. PMID: 4215937.

(13) Buchanan TM, Sulzer CR, Frix MK, Feldman RA. Brucellosis in the United States, 1960-1972. An abattoir-associated disease. PartII. Diagnostic aspects. Medicine (Baltimore). 1974 Nov; 53 (6): 415–25. doi:10.1097/00005792-197411000-00002. PMID: 4612293.

(14) Buchanan TM, Hendricks SL, Patton CM, Feldman RA. Brucellosis in the United States, 1960-1972. An abattoir-associated disease. Part III. Epidemiology and evidence for acquired immunity. Medicine (Baltimore). 1974 Nov; 53 (6): 427–39. doi:

(15) Ollé-Goig J, Canela-Soler J. An outbreak of Brucella melitensis infection by airborne transmission among laboratory workers. Am J Public Health 1987; 77:335–338.

(16) → trasmisión aérea de Rev.1.

(17) Ashford DA, di Pietra J, Lingappa J, et al. Adverse events in humans associated with accidental exposure to the livestock brucellosis vaccine RB51. Vaccine 2004; 22: 3435–3439.

(18) Wallach JC, Ferrero MC, Victoria M, et al. Occupational infection due to Brucella abortus S19 among workers involved in vaccine production in Argentina. Clin Microbiol Infect 2008; 14: 805–807.

(19) Xie J, Wang J, Li Z, et al. Ontology-based meta-analysis of animal and human adverse events associated with licensed brucellosis vaccines. Front Pharmacol 2018: 9 (article 503): 1–11.

(20) Solís J, Lorente S, Navarro E, Solera J. Detection of IgM antibrucella antibody in the absence of IgGs: a challenge for the clinical interpretation of Brucella serology. PLoS Negl Trop Dis 2014; 8 (12): e3390.

(21) Castaño MJ, Solera J. Chronic brucellosis and persistence of Brucella melitensis DNA. J Clin Microbiol 2009; 47: 2084–2089.

